# Invasive airway “Intubation” in COVID-19 patients :statistics, causes and recommendations

**DOI:** 10.1101/2021.04.08.21254439

**Authors:** Mostafa Mohammadi, Alireza Khafaee Pour Khamseh, Hesam Aldin Varpaei

**Author notes:** corespounding author.

## Abstract

**Background:** Severe COVID-19 disease could induce acute respiratory distress which is characterized by tachypnea, hypoxia, and dyspnea. Intubation and mechanical ventilation are a strategic treatment of COVID-19 distress or hypoxia.

**Methods:** We searched PubMed, Embase, and Scopus databases through April 1, 2021, to identify relevant randomized control trials, observational studies, and case series.

**Results:** 24 studies were included in this review. Studies were conducted in the USA, China, Spain, South Korea, Italy, Iran, and Brazil. Most patients were intubated in the intensive care unit. Rapid sequence induction was mostly used for intubation. ROX index might be utilized for the predictor of the necessity of intubation in COVID-19 patients. According to the previous studies the rate of intubation reported 5 to 88%. It was revealed that 1.4 - 44.5% of patients might be extubated. Yet obesity and age (elderly) are the only risk factors of delayed or difficult extubation.

**Discussion and conclusion:** Acute respiratory distress in COVID-19 patients could require endotracheal intubation and mechanical ventilation. Severe respiratory distress, loss of consciousness, and hypoxia were the most important reasons for intubation. Also, increased levels of ferritin, d-dimer, and lipase in common with hypoxia are correlated with intubation and ICU admission Mortality following intubation is reported to be 15 to 36%. Awake-prone positioning in comparison to high-flow nasal oxygen therapy did not reduce the risk of intubation and mechanical ventilation. There was no association between intubation timing and mortality of infected patients. noninvasive ventilation may have survival benefits.

## Background

Tracheal intubation is a clinical procedure to place a flexible tube into the trachea with the aim of keeping a safe airway and establish ventilation. Some circumstances might require intubation such as loss of consciousness, major surgeries, decreased oxygen saturation (hypoxemia), airway obstruction (laryngospasm), or respiratory disease such as acute respiratory distress syndrome [1,2]. Since this is an invasive and uncomfortable procedure, intubation is often performed under general anesthesia and a neuromuscular-blocking medication.

During the COVID-19 pandemic, as infected patients develop acute respiratory distress and respiratory failure, putting invasive airways might prevent disease progress [1,2,3]. Approximately 14–30% of hospitalized patients diagnosed with COVID-19 develop a severe respiratory failure requiring intensive care [4,5,6] so that roughly 3.2% of patients with COVID-19 required intubation and invasive ventilation at some point in the disease periods [7]. The need for intubation and mechanical ventilation in those who are critically ill is vary ranging from 30 to 100 percent [8].

Currently, different statistics are provided on the intubation rate of COVID-19 patients in intensive care units. There are various statistics on the intubation of patients with COVID-19. On March 4, 2020, 3.2% of infected cases in China required intubation [7]. Also, in a study conducted in New York, 12.2 to 33.1% of covid-19 patients needed intubation [4,9]. The duration of intubation is different for each patient. Wali et al examined the course of 5 COVID-19 patients who needed invasive oxygen therapy [10]. It was found that most patients were intubated in the first 2 days of admission. The duration of ventilation with an endotracheal tube was between 4 to 30 days. Intubation can be due to various reasons, like decreased oxygen saturation, respiratory distress, and Acute Respiratory Distress Syndrome [11]. In this situation, intubation at the right time is very important, and delayed intubation might cause patients’ death [12]. Xiao Lu’s studies show that mortality in intubated patients with COVID-19 is higher than in patients who did not intubate [13]. Most intubations are performed in the ICU. According to recent studies, more than 10% of COVID-19 patients in northern Italy who suffer from hypoxia were intubated in the ICU [14]. However, a unique syndrome of hypoxic COVID-19 patients has been described (labeled “the happy hypoxic”) who are mentally alert and lack of significant respiratory distress despite hypoxia that would usually prompt treatment, sometimes with profoundly low oxygen saturation [15].

The timing of intubation as well as the decision to endotracheal intubation may be unique to COVID-19 patients’ case by case [16]. The threshold for intubation may be lower in COVID-19 since the use of high-flow nasal oxygen [2] or non-invasive ventilation may potentially increase the risk of transmission to healthcare workers. Disease transmission risk of viral respiratory infections during intubation is high, and therefore early, controlled intubation may also increase the safety margin of intubation and by allowing adequate preparation time for this high-risk procedure. The application of airborne precautions is highly recommended throughout [17,18]. The most important clinical manifestations of respiratory failure in patients with COVID-19 are hypoxemia and increased work of breathing. Attention should be paid to the different effects of different oxygen therapy concentrations to avoid prolonged high-concentration oxygen therapy [19]. Due to the involvement of the alveoli in COVID-19 pneumonia, it is vital to deliver oxygen at the right concentration to maintain oxygen saturation and avoid decreased partial pressure of oxygen.

## Methods and Materials

The literature search using the following search strategy was conducted on the PubMed, Embase, and Scopus databases on April 1st, 2020 to identify eligible articles: (intubation statistics and COVID-19). The publication time was limited to 2020 onward. A total of 4,680 papers were identified by the initial search. Three reviewers independently reviewed the abstracts and full-texts. Inclusion criteria: the studies in which, among the study population, a percentage of COVID-19 infected patients (not all patients) required endotracheal intubation. Reports regarding intubation in COVID-19 which consists of the patient’s intubation rate were included in this review. The main question of this review is how many patients infected by COVID-19 were intubated? and what are associated factors?

This study was approved by the internal ethical committee of critical care, Imam Khomeini hospital complex. Ethics code number IR.TUMS.VCR.REC.1399.389.

**Figure 1.**
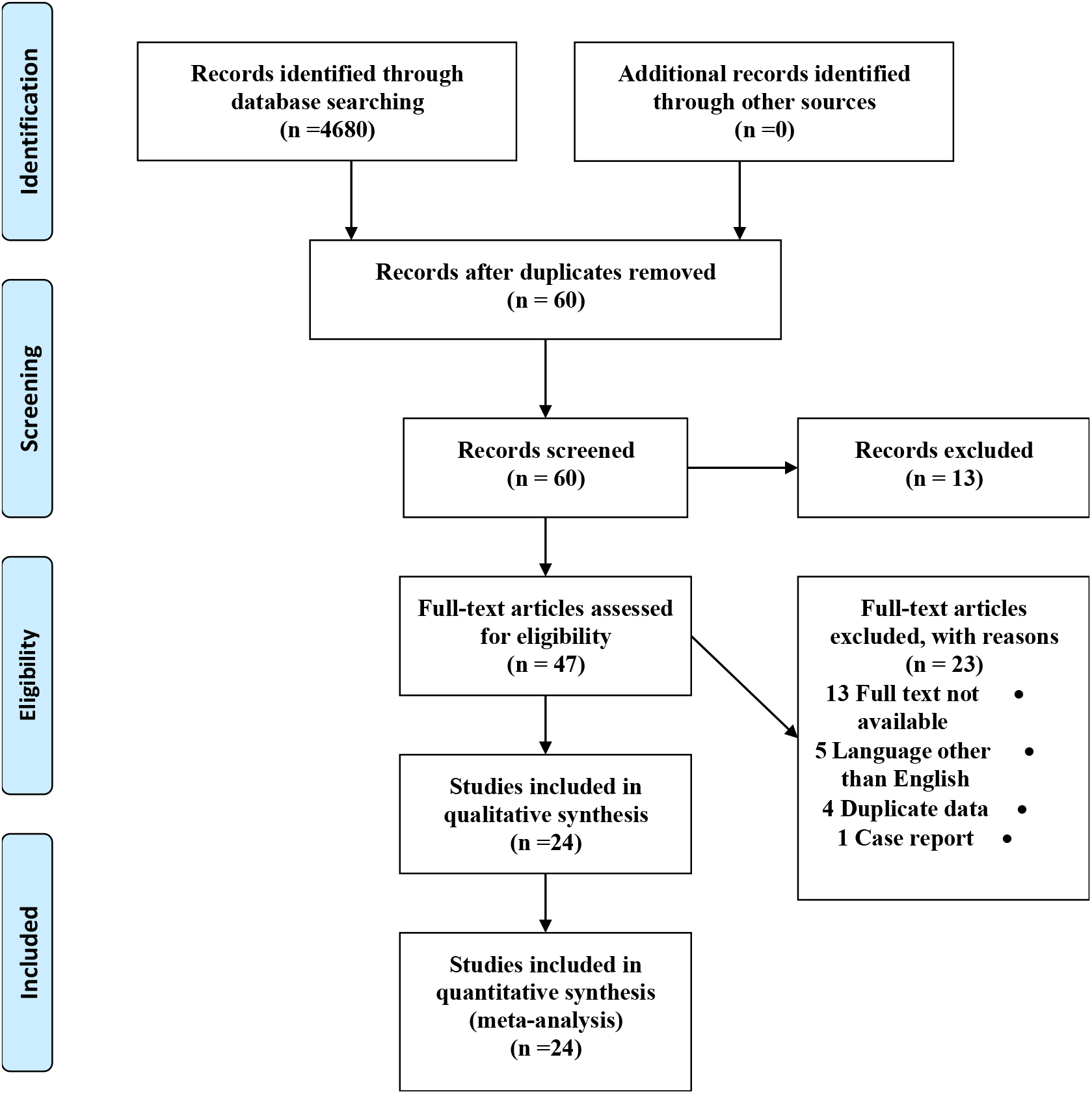
PRISMA 2009 Flow Diagram.

## Results

Overall, 24 studies regarding the intubation rate in COVID-19 patients were included in the final analysis. Most of the patients were intubated in the intensive care unit. Studies were conducted in the USA, Spain, Italy, China, South Korea, Brazil, and Iran.

**Table 1.** Statistics of COVID-19 patients Intubation

| Table 1 – Statistics of COVID-19 patients Intubation |  |  |  |  |  |  |
| --- | --- | --- | --- | --- | --- | --- |
| Sex | Age | Ward | Extubation Rate % | Intubation Rate % | Cause(s) of intubation | Country |
| 210 Male<br>128 Female | 39<br>(IQR 31- 45) | ED<sup>1</sup> | – | 8 %<br>28 out of 338 | Respiratory failure | USA, New York [20] |
| 78<br>(Not specified) | 58.4±13.7 | ICU | – | 12.82 %<br>10 out of 78 | Respiratory failure | Tabriz, Iran [21] |
| 60 % Male<br>40 % Female | 59 | ED | – | 13 % | Long term Hypoxia | USA [22] |
| Mostly Male | 50 – 81 | Oncology | – | 5 % | Hypoxia and Lung injury caused by Oxygen decreased | USA [23] |
| Mostly Male | 47 -70 | Infectious ward | – | 26.7 % | Hypoxia | USA [24] |
| 52 % (53/102)<br>Male<br>48% (49/102)<br>Female | 68<br>(IQR 61-75) | ICU | – | 57% women<br>59 out of 102 | Hypoxic respiratory failure | USA, New York [25] |

<sup>1</sup> Emergency Department

|  |  |  |  |  |  |  |
| --- | --- | --- | --- | --- | --- | --- |
| 78 % (83/108) Male<br>22 % (25/183) Female | 64<br>(IQR 57-70) | ICU | 12/822 (1.4%) | 20 %<br>163 out of 822 | – | Italy, Bologna<br>[26] |
| 137(71%) Male | 60±13.8 | – | – | 62.7 % | – | USA [27] |
| 90(53%) Male | 57±11 | ICU | – | 38.29 % | – | USA [28] |
| 53(33%) Male | 62±14.3 | ICU | – | – | ARDS | USA [29] |
| 35 (67%) Male<br>17 (33%) Female | 59.7 ±13.3 | ICU | – | 42 %<br>22 out of 52 | ARDS | China [30] |
| 1304 Male<br>287 Female | 63 (IQR 56 –<br>70) | ICU | – | 88 %<br>1150 out of 1591 | Hypoxia and<br>ARDS | Italy,<br>Lombardy [5] |
| 49% Male: PE<br>group<br>45% Male: non-PE<br>group | 62±16 PE<br>group<br>59±15 non-PE<br>group | ICU | – | 65 % in the PE<sup>2</sup><br>group<br>67 % in the non-PE<br>group | Hypoxia<br>and PE | [58] |
| No data available | No data available | ICU | 44.5%<br>842 out of 1890 | 16.4 %<br>1890 out of 11493 | - | Spain [35] |
| 9 Male<br>7 Female | 58<br>(IQR,47 – 68) | ICU | - | 56.25 %<br>9 out of 16 | Respiratory failure | Wuhan, China [13] |
| No data available | No data available | ICU | - | 41.20 %<br>82 out of 199 | Acute respiratory failure | Spain and Andorra [33] |
| 28 Male<br>19 Female | 70 years<br>(IQR, 63–77) | ICU | - | 48.9 %<br>23 out of 47 (early intubation*)<br>34 %<br>16 out of 47 (late intubation**) | ARDS | Daegu, South Korea [34] |
| 88 Male<br>50 Female | 55 cases ≤ 60 years<br>83 > 60 years | ED, ICU and other wards | 56.5 %<br>78 out of 138 | 24.8 %<br>138 out of 486 | Decreased Oxygen saturation, Shortness of breath | Chicago, USA [35] |
| 92 Male<br>38 Female | 64.5 years<br>(IQR, 51.7–73.6) | – | 33.1 %<br>43 out of 130 | 33.1 %<br>130 out of 393 | Dyspnea | New York, USA [9] |
| 25 Male<br>5 Female | 65.5 years<br>(IQR, 57–71) | – | 20 %<br>6 out of 30 | 23.25 %<br>30 out of 129 | Hypoxemia | New Jersey, USA [36] |
| No data available | 65 years<br>(IQR, 53–77) | Various wards | – | 10.6 %<br>467 out of 4389 | Decreased Oxygen saturation, Hypoxia | New York, USA [37] |
| 37 Male<br>45 Female | No data<br>available | – | – | 33.75 %<br>27 out of 82 | Hypoxia | Chicago, USA<br>[38] |
| 112 Male<br>54 Female | 58.1±14.1 | ED | – | 58 % (prone<br>positioning)<br>33 out of 57<br>49 % (No Prone)<br>53 out of 109 | Hypoxia,<br>tachypnea | Sao Paulo,<br>Brazil [39] |
| 129 Male<br>93 Female | 69.5<br>(IQR, 62–78) | Various<br>wards | – | 41 % (Intubation<br>First)<br>91 out of 222<br>20 % (NIV to<br>intubation)<br>44 out of 222 | ARDS,<br>Hypoxia,<br>Altered<br>Mental Status | New York,<br>USA [40] |

\* Early intubation: intubation as soon as ARDS diagnosis
\*\* Late intubation: not initially intubated, but subsequently required intubation during follow-up

Various statistics of intubation rates in COVID-19 patients are reported ranging from 5 – 88 %. This difference in statistics can be due to differences in sample size, study environment (wards), and intubation criteria [22,25]. Rapid sequence induction (RSI) or modified RSI was mostly used for intubation [41]. Propofol was used mostly in all patients often combined with other sedative agents like Rocuronium, Sufentanil, and Midazolam. Studies showed that factors affecting COVID-19 severity (such as underlying disease, age more than 50, smoking, body mass index, and comorbidity condition) could exacerbate a patient’s condition and accelerate the necessity for endotracheal intubation [23,25]. The most important reasons that require endotracheal intubation are hypoxia, respiratory distress, loss of consciousness [13,20,21]. In a study by Xu, et al. it was found that the intubation of critically ill patients with COVID-19 is mostly due to respiratory failure, but there is also a small number of patients who undergo intubation due to secondary acute heart failure or airway obstruction [42].

Some studies revealed that due to the limited capacity of intensive care units during the COVID-19 pandemic [43], tracheostomy seems to be a suitable solution for patients to get off the mechanical ventilation [44], reducing the respiratory effort in patients with limited pulmonary reserves, shortening the dead space and enabling the suctioning of accumulated mucous [32].

Despite the rarity of available studies regarding extubation in COVID-19 patients’, some evidences revealed the extubation rate ranging 1.4 % - 44.5% [26,32].

As this study is a retrospective study, the number of studies included in this investigation is relatively small, and further large-scale prospective studies are needed to confirm our findings.

## Discussion and Conclussion

SARS-CoV-2 is the coronavirus responsible for the COVID-19 pandemic of 2020. COVID-19 might induce severe respiratory distress. Acute respiratory distress in COVID-19 patients could require endotracheal intubation and mechanical ventilation. Proper intubation and management of patients with coronavirus are integral parts of critically ill patient’s management. In confirmation, a study stated that of the patients who died only ∼ 25% received invasive mechanical ventilation (intubated) or ECMO [45]. It seems that lack of ventilator or delayed intubation might be one of the causes of deterioration disease in COVID-19 patients.

A study suggested that [46], less than a third of patients with coronavirus, might benefit from high-flow nasal cannula therapy, noninvasive ventilation, and awake prone position. Patients who have extubated might also benefit from this treatment strategy. The use of awake prone positioning as a supplement therapy to high-flow nasal oxygen therapy did not reduce the risk of intubation and mechanical ventilation [47]. However, it could cause delayed in intubation [33]. It seems that there are no significant differences in mortality rate between the early group and never intubated patients [34].

According to the necessity of intubation, specialists suggested the use of esophageal manometry (as a surrogate of pleural pressure) and consider intubation when pressure swings exceed 15 cm H2O identifying risk of self-inflicted lung injury [48]. Experience-based recommendations stated; once the patient’s non-invasive ventilation failed, elective intubation should be preferred to minimize clinical risks such as contamination of medical staff [49]. Adequate application of personal protective equipment would reduce the risk of nosocomial infections for proceduralists [50].

Several factors might be associated with intubation in COVID-19 patients. Elderly patients, male gender and background diseases particularly hypertension and diabetes are more exposed to the risk of intubation [1,35,50]. Increased levels of ferritin, d-dimer, and lipase in common with hypoxia are correlated with intubation and ICU admission [36,38]. It was found that anticoagulation is associated with lower intubation [37]. The result of a study suggests that the ROX index is a noninvasive and outstanding predictor for the necessity of intubation in COVID-19 patients [51]. The ROX index is the ratio of oxygen saturation as measured by pulse oximetry/FiO2 to respiratory rate, it is assessed as a predictor of the need to intubate in patients who received high flow nasal cannula oxygen therapy [52].

Although appropriate positioning is an essential part of successful intubation, a study stated that awake prone positioning may not correlate with intubation rates [39]. Furthermore, there was no significant change in terms of PaO2 before Prone Positioning and PaO2 after resupination [53].

It seems that noninvasive ventilation (NIV) may have survival benefits so that patients who received NIV have lower mortality than patients who did not receive NIV and were intubated [40]. A study by Solaimanzadeh suggested that some calcium channel blocker agents like Nifedipine and Amlodipine may remarkably meliorate mortality and reduce the risk of intubation and the need for mechanical ventilation in Elder COVID-19 patients [54].

According to the studies the intubation rate reported 5 to 88 %. In the review study by da Silva CM et al, intubation need vary from 2, 3, 4, to 42% and 47% [55]. Besides, Cardona et al reported that the intubation rate was 28% among hypoxic COVID-19 patients [56]. This discrepancy in statistics may be due to the variety of study population, study environment, or intubation criteria. However, there are a few pieces of evidence regarding extubation, therefore; more investigations are required to determine intubation outcomes and extubation. A study stated that obesity and age are the only risk factors of delayed or difficult extubation [35].

In terms of mortality subsequent of COVID-19 patients’ intubation, it was reported vary from 15.2% [35], 23.1% [21], 24.5% [4], to 36.0% [57]. Hernandez-Romieu et al [47] found no association between time to intubation and mortality of infected patients.

Due to the lack of definitive treatment for COVID-19, and the empirical treatments based on research, it seems that a comprehensive intubation algorithm in COVID-19 patients is extensively required.

The findings of this study are a review of intubation rates based on previous studies. These statistics are preliminary and more research is needed to examine the correlation and the factors affecting the intubation of patients.

## Recommendations

According to studies and experience, we hereby offer recommendations that might improve the quality of care.

Utilize full personal protective equipment to prevent transmission.

It is better than the most experienced specialist intubates patients. (Perfectly first attempt)

Noninvasive ventilation may correct oxygenation and have survival benefits. It can be considered for patients who tolerate it.

Rapid sequence induction in combination with video laryngoscopy enabled swift intubation [30].

Necessary equipment should be available prior to intubation procedure (monitoring, intravenous access, resuscitator medications, ventilator, and suction) [61]

Consider calcium channel blocker agents (particularly Nifedipine and Amlodipine) for hypertensive patients [60]. (if applicable)

## Data Availability

All data were collected from PubMed, Embase, and Scopus and google scholar databases.All data (articles which mentioned in references) are available online.

## Acknowledgement

We hereby thanks all healthcare providers (hospital staff, nurses, doctors) around the world. We are grateful to the esteemed faculty members of Tehran University of medical sciences, the anesthesiology, and critical care department. #thanks_heros

